# Heart rate and heart rate variability in patients with chronic inflammatory joint disease: The role of pain duration and the insular cortex

**DOI:** 10.1101/2021.01.14.21249826

**Authors:** Leona Katharin Buschmann, Melanie Spindler, Peter Sörös, Carsten Bantel

## Abstract

Chronic inflammatory joint diseases (CIJD) have been linked to increased cardiovascular morbidity and mortality. A decisive reason could be a dysregulation of the autonomic nervous system (ANS), which is responsible for the control of cardiovascular function. So far, the cause of changes in ANS functions remains elusive. In this study, we investigate the role of chronic pain and the insular cortex in autonomic control of cardiac functioning in patients with CIJD. We studied the sympathetic and parasympathetic branch of the ANS through the assessment of heart rate and heart rate variability (HRV) at rest and under cognitive stimulation. Furthermore, we investigated insular cortex volume by performing surface-based brain morphometry with FreeSurfer. For this study, 22 individual age- and sex-matched pairs for the magnetic resonance imaging analyses and 14 for the HRV analyses were recruited. Pain duration was negatively correlated with the resting heart rate in patients with chronic inflammatory joint diseases (n = 19). In a multiple linear regression model including only CIJD patients with HR at rest as a dependent variable, we found a significant positive relationship between HR at rest and the volume of the left insular cortex and a significant negative relationship between HR at rest and the volume of the right insular cortex. However, we found no significant differences in HRV parameters or insular cortex volumes between both groups. In this study we provide evidence to suggest insular cortex involvement in the process of ANS changes due to chronic pain in CIJD patients.

## Introduction

Chronic inflammatory joint diseases (CIJD), such as rheumatoid arthritis (RA) and ankylosing spondylitis, are commonly associated with disability and pain [37]. CIJDs have also been linked to increased cardiovascular morbidity and mortality, which cannot be attributed to typical risk factors, such as smoking or hypertension [14; 22; 44; 68; 81]. Instead, a dysregulation of the autonomic nervous system (ANS) has been suggested as a potential underlying mechanism. This notion is based on an estimate of approximately 60% of patients with RA showing signs of ANS dysfunction [5].

The function of the ANS can be assessed by measuring heart rate and calculating heart rate variability (HRV), i.e. the variation of the length of the R-to-R-wave time interval [34; 63]. In patients with rheumatic diseases, a reduction of HRV has been observed, indicating reduced cardiac adaptability [5; 24; 36]. However, the reasons for these changes in ANS function in CIJD remain elusive. Based on the results of a study by Dunn and Croft from 2006, who showed the prognosis of low back pain was dependent on the duration of pain, it could be possible that changes in HRV might be associated with pain duration [12].

The ANS is controlled by centers in the brainstem, the hypothalamus, and, ultimately, by several cortical regions, including the insula, anterior cingulate gyrus, and medial frontal cortex [9; 55]. The insula is considered a crucial region for polymodal sensory, cognitive, and affective processing [23]. In addition to the processing function of somatosensory [72; 73], peripheral inflammatory [40], and nociceptive stimuli [29], the insular cortex participates in the control of arbitrary and involuntary movements, such as speech production [4] and swallowing [71]. Oppenheimer et al. were first to demonstrate changes in cardiac function after electrical stimulation of the left and right insula in humans [51]. Corroborating these results, several studies reported cardiac dysfunction following insula lesions due to stroke, tumors, or surgical resections [16; 47; 65; 70].

Wartolowska and colleagues used surface-based and voxel-based morphometry as well as segmentations of subcortical nuclei to investigate morphological changes in the brains of patients with rheumatoid arthritis. This study found an increase in in gray matter in the basal ganglia of the patient group, mainly in the nucleus accumbens and caudate nucleus, without significant differences in cortical gray matter [80]. Similarly to what has been described for other chronic pain conditions such as chronic back and limb pain [8; 56], complex regional pain syndrome [30], migraine [58], chronic tension type-headache [59], and fibromyalgia [41], patients with CIJD might also display structural changes in cortical regions that are involved in the regulation of the ANS, especially in the insular cortex.

The aims of the present study hence were a) to evaluate differences in HRV at rest and under stimulation between the CIJD and control groups, b) to test whether pain duration might be associated with changes in heart rate and heart rate variability in patients with chronic inflammatory joint diseases, and c) whether altered HRV is associated with structural changes in ANS-controlling cortical areas such as the insular cortex.

## Methods

The study was approved by the Medical Research Ethics Board of Carl von Ossietzky University of Oldenburg, Germany (#2017-059) and was preregistered with the German Clinical Trials Register (https://www.drks.de; DRKS00012791). All participants gave written informed consent prior to participating in this study. Participants (healthy controls: C, patients with chronic inflammatory joint diseases: CIJD) were recruited between July 2017 and March 2019. All measurements were performed at the Neuroimaging Unit, School of Medicine and Health Sciences, Carl von Ossietzky University of Oldenburg. All data were collected as a part of a larger study, which, in addition to the data evaluated here, included functional magnetic resonance imaging (fMRI) and behavioral tests [69; 74].

### Participants

Patients aged between 18 and 75 years were recruited at a specialized rheumatological outpatient clinic and with the help of a support group for ankylosing spondylitis patients in Oldenburg. Controls were recruited by advertisements in a local newspaper and announcements on the university’s website. Participants had to be fluent in German and right-handed. Patients needed to suffer from a chronic inflammatory joint disease that caused chronic pain for at least one year while controls had to be free of CIJD and chronic pain in general. Controls and CIJD were matched for sex and age (± 5 years).

Pregnant women and participants with contraindications for MRI were excluded, in addition to participants with conditions that could have had an influence on HRV or cerebral functions: previous heart surgeries; heart failure stage > 2 of the New York Heart Association’s classification [53]; peripheral artery diseases type 2 - 4 according to Fontaine [3]; neurological diseases (e.g. stroke, multiple sclerosis, myasthenia gravis, known cerebrovascular disease); active cancer; chronic liver diseases (hepatitis or liver failure); renal failure requiring dialysis; severe asthma and severe chronic obstructive pulmonary disease (COPD); poorly adjusted thyroid diseases; infections (tuberculosis, human immunodeficiency virus); psychiatric disorders (schizophrenia, severe depression, borderline personality disorder) and dyscalculia. Finally, patients with medications that might have influenced the conduction system of the heart or/and active or previous alcohol or drug abuse were also excluded.

### Experimental procedure

First, each participant completed a questionnaire including information about demographic and clinical characteristics, such as pain, co-morbidities, and suitability for MRI-examinations. Next, structural and functional MRI was performed while the measurements of HRV at rest and under cognitive stimulation were carried out.

### Data acquisition

Information about the diagnosis, pain intensity, duration of pain, and medications were collected in the CIJD group. Pain intensity was assessed using an 11-point numerical rating scale (NRS, 0 = no pain and 10 = worst pain imaginable) [12]. Photoplethysmographic pulse oximetry was used to determine HRV. Data recording was performed using the integrated Siemens Physiological Monitoring Unit of the body scanner (MAGNETOM Prisma, Siemens, Erlangen, Germany). In short, an infrared emitter was placed on the left index finger. The pulse curve was recorded at 50 Hz and heart rates, as well as the length of the intervals between heartbeats, were determined. All signals were transmitted wirelessly to the scanner and saved for later analysis.

MR images were acquired by a research-dedicated 3T whole-body scanner (MAGNETOM Prisma, Siemens, Erlangen, Germany) with a 64-channel head/neck coil. T1-weighted images were obtained with a magnetization prepared rapid gradient echo sequence (MP-RAGE) with the following parameters: voxel size: 0.75 × 0.75 × 0.75 mm, 224 sagittal slices, repetition time (TR): 2000 ms, echo time (TE): 2.07 ms, inversion time (TI): 952 ms, anterior to posterior phase encoding direction, flip-angle: 9°, in-plane acceleration with an acceleration factor of 2 (GRAPPA), and an acquisition time of 6:16 minutes.

### Experimental paradigm

Three functional MRI experiments were performed: an arithmetic experiment with two subtraction tasks, a number line identification experiment, and a resting-state measurement. Here, the HRV signals recorded during the arithmetic experiment and the resting-state measurement were analyzed as a marker of autonomic function [76]. The analysis of task-based and resting-state fMRI data will be presented in another publication.

As in the Trier social stress test, serial subtraction was employed to stimulate a cardiac response [39]. The arithmetic experiment consisted of four conditions: (1) simple serial subtraction (e.g., starting with 99 - 7), (2) difficult serial subtraction (e.g., starting with 173-13), (3) continuously reading aloud a number presented on the screen, and (4) a rest condition, fixating a cross on the screen. Every condition lasted 20 s and was repeated with different numbers five times during the experiment. For the subtraction conditions, the participants were instructed to perform serial subtractions as quickly as possible and say out loud every result. Finally, after a short break of less than a minute, a resting state measurement took place. Short breaks between the tasks were sufficient, as the HRV should return rapidly to a normal level after temporary stimulation [1].

### Coupling MRI tasks and pulse oximetry measurements

The Physiological Artifact Removal Tool (PART, https://www.mccauslandcenter.sc.edu/crnl/tools/part) was used to link the starting points of the MRI tasks and pulse oximetry measurements. The MRI measurements included 205 volumes each in the arithmetic task and resting state. For the analysis of the pulse oximetry measurements, the first 4 volumes were deleted. This led to a duration of 6 minutes and 42 seconds for each HRV measurement.

### Analysis of heart rate variability

Data processing and calculation of the HRV via pulse oximetry measurements were performed with Kubios HRV standard (Version 3.2.0, http://www.kubios.com/), which is an established software for assessing ANS function [75]. Artifacts such as extra, missing, or ectopic beats can cause considerable distortion of HRV results [11]. Therefore, ‘threshold-based correction’ was employed and preset to ‘low’. The correction algorithm detected and corrected every beat-to-beat interval that was 0.35 seconds longer or shorter compared to the local average calculated using an internal software algorithm. Corrected data were divided into time and frequency domain parameters. Time-domain parameters included the heart rate (HR) in beats per minute (bpm), and the standard deviation of all normal-to-normal beat intervals (SDNN) in ms, which reflects the variation within the R-waves intervals. The frequency data were divided into low frequency (LF) and high frequency (HF) bands. The absolute power of the different frequency bands was specified in ms^2^/Hz. The frequency ranges for the absolute power measurements were as follows: LF = 0.04 - 0.15 Hz and HF = 0.15 - 0.4 Hz [75]. LF and HF were used to characterize the response of the ANS. The HF band is thought to reflect the activity of the cardiac parasympathetic function [46; 63]. The physiological meaning of the LF is controversial. Partly representations of the sympathetic and parasympathetic nervous system, separate sections of the parasympathetic branch of the ANS, or the oscillations of the baroreflex, are discussed [31; 54; 63]. HR, SDNN, LF, and HF are some of the most used parameters for HRV analysis [49; 63].

### Analysis of MR-images

The image analysis, performed with FreeSurfer (Version 6.0.0, released January 2017, http://surfer.nmr.mgh.harvard.edu) on Mac OS high sierra (Version 10.13.6), started with the extraction of the cortical (pial) surface. Due to the hybrid watershed/surface deformation procedure, non-brain tissues were removed [60]. Additionally, segmentation of gray and white matter and intensity normalization was performed [26; 66]. The surfaces were constructed as a grid of triangles, placed in areas with the greatest intensity shift. The intensity shift was caused by the transition between different tissues [20]. Next, the software modeled the surface between gray and white matter and the outer-pial surface, which represented the boundary of gray matter to cerebrospinal fluid. The final step was the automated topological correction to fix segmentation errors and produce a closed sphere [25; 61]. Regardless of the MRI scanner and field strength, the morphometric test-retest reliability is high [33]. The quality of the created surfaces and subcortical structures was visually checked for each participant.

### Cortical volume analysis

FreeSurfer performed an automatic reconstruction of the brain’s surface. The segmentation procedure and the parcellation of the cerebral cortex were one of the steps of the command `recon-all`, which included the extraction of the white matter and other structures, a vertex-wise reconstruction of the surfaces, smoothing, inflation, and localization of topological defects. In the end, possible tracked defects were automatically fixed. The system used the probability of the localizations of subcortical structures [26; 27]. In the process of cortical parcellation, the units were assigned according to gyral and sulcal structures. Data evaluation was based on the atlas by Desikan et al. [21; 28]. The regions of interest (ROI) for this study were the insulae as processing centers of the ANS in the left and right hemispheres. The cortical brain data were normalized using the estimated total intracranial volume (eTIV) generated by FreeSurfer [13; 45].

### Statistical Analysis

All statistical analyses were performed using SPSS 25 (IBM, Ehningen, Germany). Data were analyzed for normal distribution employing Shapiro-Wilk tests. SDNN, LF, and HF were logarithmically transformed to approximate normal distribution. Demographic data were first analyzed descriptively. To determine possible differences between the demographic data of the groups, independent samples t-tests and chi-square tests were used. Repeated measure analysis of variance (ANOVA) was employed to evaluate possible differences between the HRV parameters in the different measurements and between the groups. The effect size *η*^2^ was calculated and interpreted according to Cohen (< 0.01 = small, 0.06 = medium and > 0.14 = large effect) [17].

Bivariate Pearson’s correlation analyses between HRV at rest and under stimulation, together with the pain duration in years were performed. The obtained correlation coefficients were interpreted according Cohen as small (r = 0.1), medium (r = 0.3), or large (r = 0.5) [17]. Independent samples t-tests of the left and right insular cortex volume were performed between groups. Furthermore, four linear regression models were used to test the effect of independent influences. SDNN and HR at rest were included in the first model as dependent variables while group, age, sex, and volume of the normalized insulae in both hemispheres were defined as independent variables. HR and SDNN at rest were selected for initial analysis. Finally, a second regression with only CIJD was computed with SDNN and HR at rest as dependent variables. The model corresponded to the regression model described above, only that the variable age was exchanged for pain duration and the variable group was deleted. Results were considered statistically significant if p < 0.05.

## Results

### Characteristics of participants

Figure 1 summarizes the recruitment process; 47 participants were initially recruited (23 controls, 24 CIJD patients). One CIJD patient was excluded due to limited compliance during the MRI acquisition and one control participant due to beta-blocker intake. Five control and four CIJD participants were excluded from HRV analysis because of incomplete pulse recordings or poor data quality. Participants were matched on an individual basis: For every CIJD patient, a control participant of the same sex and similar age (± 5 years) was recruited. For 22 CIJD patients and 22 matched controls, MRI data were available. For 19 corticoids previously. No control participant had ever taken glucocorticoids. Antidepressants had been taken previously by one person in each group. One CIJD patient was taking a low-dose tetracyclic antidepressant at the time of the study and none control participant. No included participant was on antihypertensive medication that could have influenced the autonomic nervous system.

**Fig. 1.**
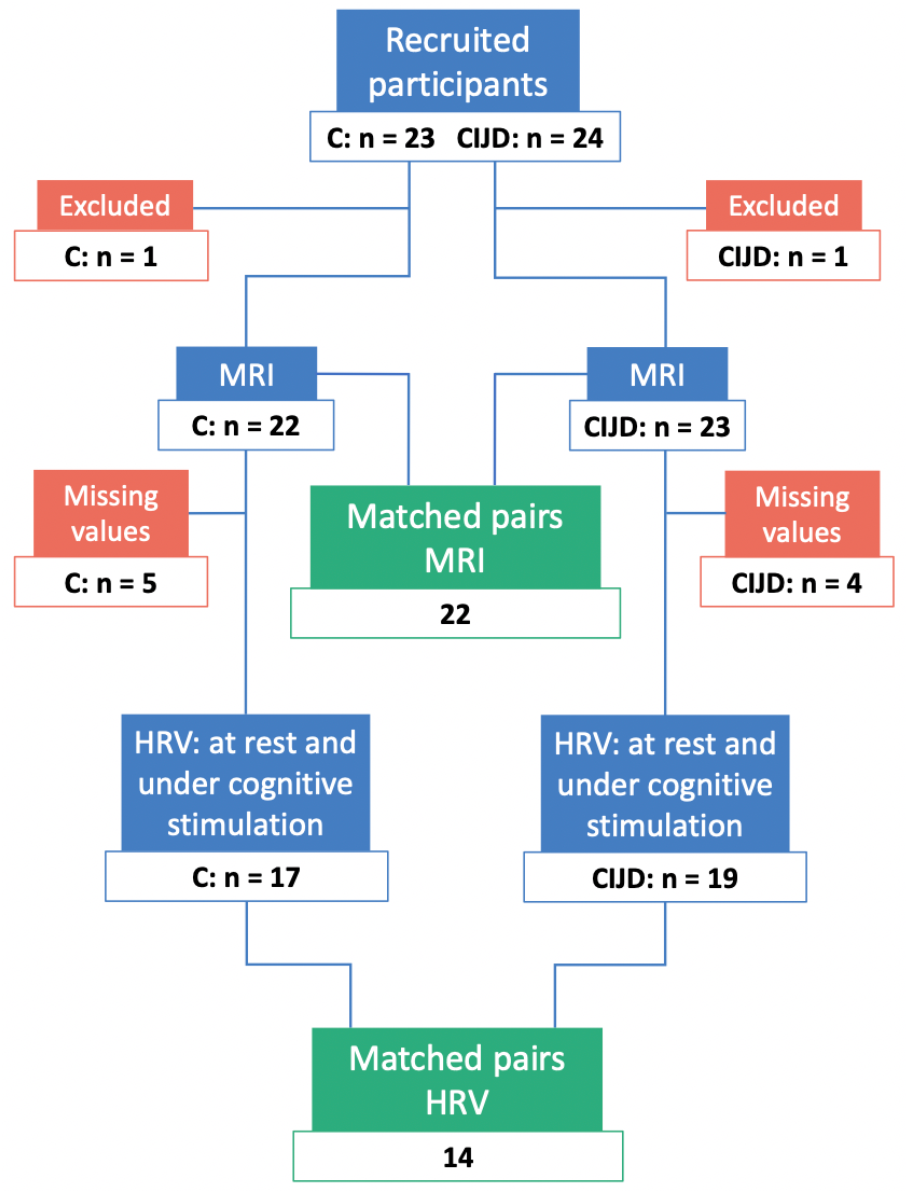
Flow diagram of participant inclusion. The diagram illustrates the participant in- and exclusion process. Blue boxes: included participants; red boxes: excluded participants. Green boxes: sex and age (± 5 years) matched pairs. Blue boxes with HRV-label: number of participants with HRV measurements under resting conditions and cognitive stimulation. C: controls; CIJD: patients with chronic inflammatory joint diseases; HRV: heart rate variability; MRI: magnetic resonance imaging

CIJD patients and 17 controls, complete pulse recordings at rest and under cognitive stimulation of sufficient quality were available, resulting in 14 matched pairs.

Table 1 summarizes the demographic and clinical characteristics of the patient and control groups. There were no significant differences between groups regarding age, smoking status, or formal education. No participants were taking opioids, benzodiazepines, or anticonvulsants at the time of the study. Seven CIJD patients were taking glucocorticoids at the time of the study; four CIJD patients had taken glucocorticoids previously. No control participant had ever taken glucocorticoids. Antidepressants had been taken previously by one person in each group. One CIJD patient was taking a low-dose tetracyclic antidepressant at the time of the study and none control participant. No included participant was on antihypertensive medication that could have influenced the autonomic nervous system.

**Table 1.** Characteristics of participants

|  | Controls | CIJD | p |
| --- | --- | --- | --- |
| Sample size, n | 22 | 22 |  |
| Gender (m/f) | 6/16 | 6/16 |  |
| Age (years), mean (SD) | 50.9 ($\pm 10.6$) | 51.3 ($\pm 10.2$) | 0.896 |
| Smoking (yes/no/earlier) | 17 / 2 / 3 | 14 / 3 / 5 | 0.609 |
| Education* (median) | 3 | 2 | 0.550 |
| Chronic inflammatory joint diseases: |  |  |  |
| – Rheumatoid arthritis; n |  | 9 |  |
| – Ankylosing spondylitis; n |  | 9 |  |
| – Psoriatic arthropathy; n |  | 3 |  |
| – CREST syndrome**; n |  | 1 |  |
| Pain duration (yrs), median (min/max) |  | 10 (1 / 44) |  |
| Pain intensity, median (min/max)*** |  | 3.5 (0 / 7) |  |

SD: Standard deviation; CIJD: patients with chronic inflammatory joint diseases.
\* The level of education was classified according to the achieved academic degree: 0 = no degree, 1 = secondary school, 2 = middle school, 3 = High school diploma, 4 = university/ college. \*\* CREST = calcinosis, Raynaud's phenomenon, esophageal motility abnormalities, sclerodactyly and telangiectasia. \*\*\* 11-NRS = 11-point numerical rating scale (0 = no pain to 10 = worst pain imaginable), on the day of testing under usual medication

### Heart rate and HRV

Table 2 displays the results of the repeated measurements’ ANOVA for HR, SDNN, LF, and HF of the patient and control groups at rest and during cognitive stimulation. In both, controls and patients with CIJD, cognitive stimulation resulted in a significant increase in HR (C: + 10.68%, CIJD: + 10.73%) with a large effect size and SDNN (C: + 6.11%; CIJD: + 8.08%) with a medium effect size. Importantly, there were no significant effects between groups. Furthermore, the LF increase in CIJD patients (C: + 2.23%, CIJD: + 12.26%) was significant with a medium effect size and no differences in the HF parameters between groups.

**Table 2.** Results of the four repeated measurements ANOVAS assessing the influence of the condition and group on HRV parameters

| | F | df | p | $\eta^2$ |
| --- | --- | --- | --- | --- |
| **Heart rate** |  |  |  |  |
| Condition | 40.950 | 1, 26 | <0.001* | 0.612 |
| Group | 0.182 | 1, 26 | 0.673 | 0.007 |
| Interaction | 0.005 | 1, 26 | 0.942 | <0.001 |
| **SDNN** |  |  |  |  |
| Condition | 12.090 | 1, 26 | 0.002* | 0.317 |
| Group | 0.377 | 1, 26 | 0.545 | 0.014 |
| Interaction | 0.254 | 1, 26 | 0.619 | 0.010 |
| **Low frequency power** |  |  |  |  |
| Condition | 10.362 | 1, 26 | 0.003* | 0.285 |
| Group | 0.237 | 1, 26 | 0.631 | 0.009 |
| Interaction | 4.851 | 1, 26 | 0.037* | 0.157 |
| **High frequency power** |  |  |  |  |
| Condition | 3.140 | 1, 26 | 0.088 | 0.108 |
| Group | 0.032 | 1, 26 | 0.859 | 0.001 |
| Interaction | 0.442 | 1, 26 | 0.512 | 0.017 |

CIJD patients demonstrated a significant negative correlation between heart rate at rest and pain duration (r = −0.627, p = 0.004, Figure 3A). During cognitive stimulation, there was no correlation between the heart rate and pain duration (r =-0.249, p = 0.277, Figure 3B). There were no correlations between SDNN, LF, or HF and pain duration, neither at rest nor during cognitive stimulation.

**Fig. 2.**
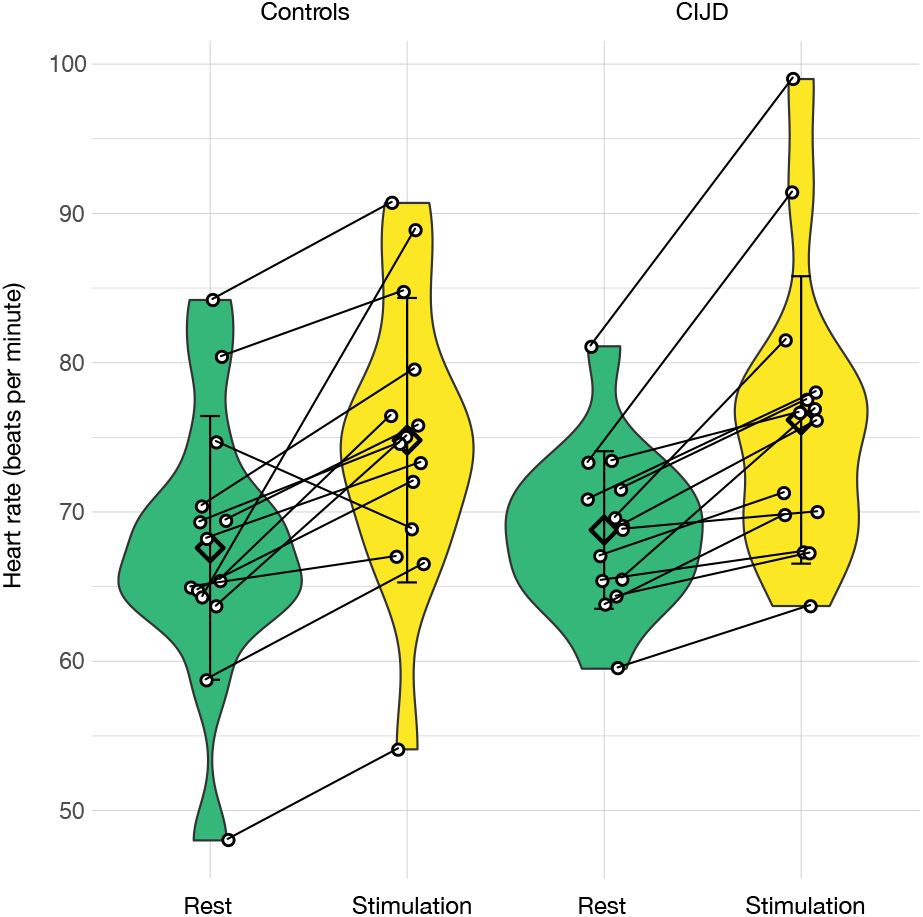
Violin plot of heart rate at rest and under cognitive stimulation. Controls (n= 14) are shown on the left and patients with chronic inflammatory joint diseases (CIJD; n = 14) on the right. Diamonds represent the mean and error bars represent the standard deviation. Green: heart rate (HR) at rest; yellow: heart rate under cognitive stimulation. The ANOVA with repeated measurement showed an increase of HR during stimulation within both groups (condition: p < 0.001, *η*^2^ = 0.612). However, there were no differences in HR at rest and under stimulation between matched groups (group: p = 0.673, *η*^2^ = 0.007).

**Fig. 3.**
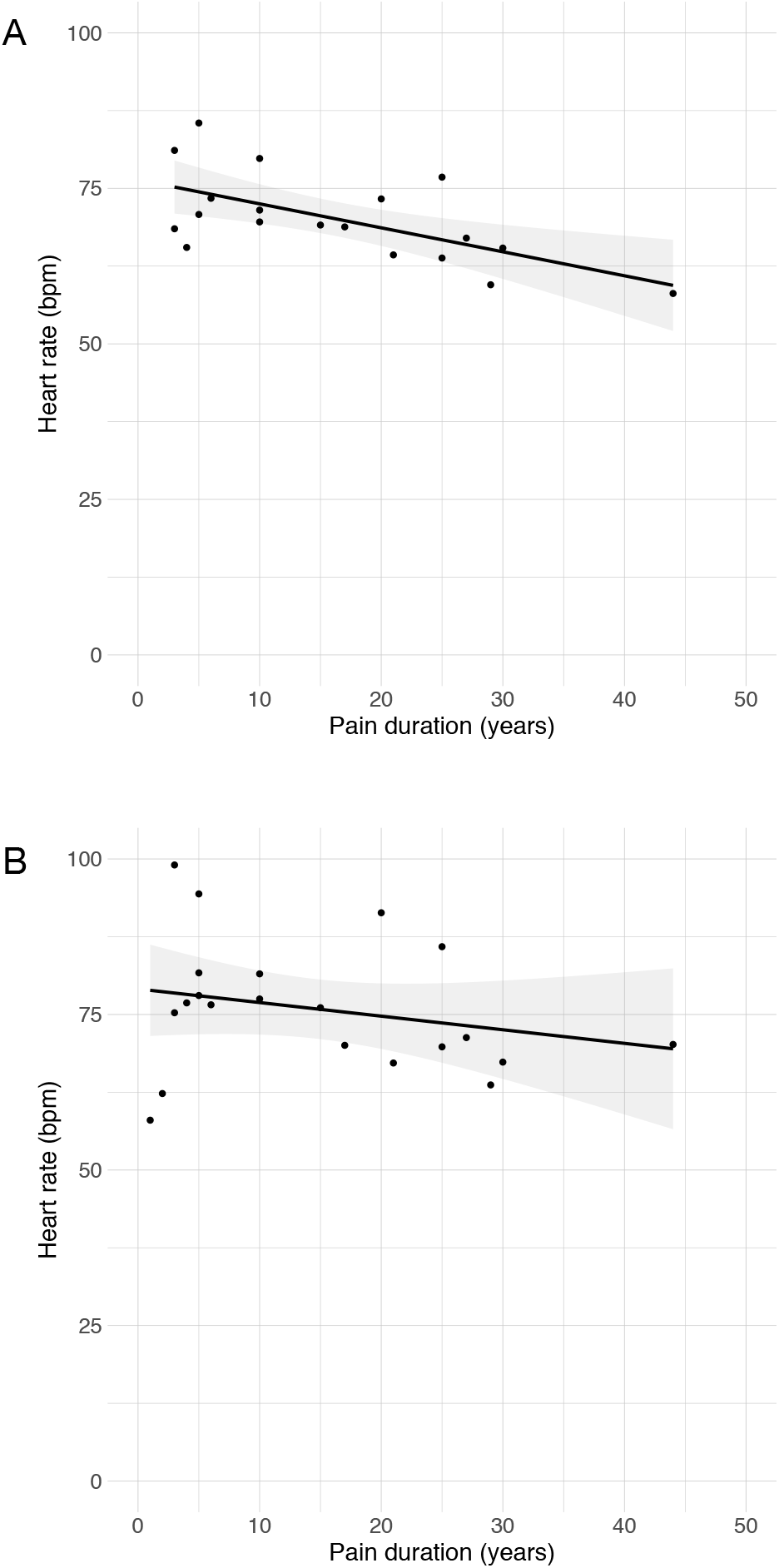
Correlation between heart rate and pain duration in patients with CIJD. **A**) Pearson’s correlation indicates a strong significant association between heart rate at rest and pain duration (p = 0.004, r = −0.627). **B**) No significant correlation between heart rate under stimulation and pain duration (p = 0.277, r = −0.249). The gray area is the 95% confidence interval with the regression line displayed.

### Structural imaging

Employing independent t-tests of insula volume with eTIV normalization (C = 22, CIJD = 22), we found no significant differences between the matched groups in the left (t(42) = 0.214, p = 0.832) and the right insula (t(42) = 0.165, p = 0.870).

The general linear regression model including group (C = 18, CIJD = 19) and HR as dependent variables showed no significant differences (p = 0.349, adjusted R^2^ = 0.022). The SDNN model including group, age, sex, and insula volume in both hemispheres showed significant results and explained 23% of the variability (p = 0.022). However, the only relevant variable was sex. The second subgroup regression model (n = 19) was used to predict HR with pain duration, sex, and volume of the insular cortices in both hemispheres. The subgroup regression explained 66% of the HR and 39% for the SDNN variability (HR: p < 0.001, SDNN: p = 0.021). Tables 3 and 4 show the significant results of both regression analyses.

**Table 3.** Linear regression model for CIJD patients with heart rate at rest as dependent variable

| Predictors | Coefficient | SE | Beta | T | p |
| --- | --- | --- | --- | --- | --- |
| Constant | 65.33 | 21.95 |  | 2.98 | 0.009* |
| Pain duration | -0.42 | 0.12 | -0.56 | -3.66 | 0.002* |
| Sex | 6.57 | 2.75 | 0.34 | 2.39 | 0.030* |
| Left Insula | 11919.12 | 4816.80 | 0.38 | 2.47 | 0.026* |
| Right Insula | -10209.36 | 3937.89 | -0.45 | -2.59 | 0.020* |

\* $p < 0.05$

**Table 4.** Linear regression model for CIJD patients with SDNN at rest as dependent variable

| Predictors | Coefficient | SE | Beta | T | p |
| --- | --- | --- | --- | --- | --- |
| Constant | 2.33 | 0.64 |  | 3.66 | 0.002* |
| Pain duration | 0.01 | 0.003 | 0.37 | 1.79 | 0.093 |
| Sex | -0.28 | 0.08 | -0.66 | -3.52 | 0.003* |
| Left Insula | -149.14 | 139.82 | -0.22 | -1.07 | 0.303 |
| Right Insula | -1.26 | 114.31 | -0.003 | -0.01 | 0.991 |

\* $p < 0.05$

## Discussion

Our results provide evidence for an inverse correlation between HR and pain duration in patients with chronic inflammatory joint disease at rest. Moreover, both healthy volunteers and patients with CIJD showed a similar adequate cardiovascular response during cognitive stimulation (serial subtraction). Finally, our data suggest that structural changes in the insular cortices are related to changes in heart rate in CIJD patients.

### Heart rate and heart rate variability

The present data provide evidence for a negative association between HR and pain duration with a large effect size (r = −0.627) at rest. To the best of our knowledge, this is the first report of an association between resting HR and pain duration in patients with CIJD. This finding may be explained by an increase in the parasympathetic or a decrease in the sympathetic outflow of the ANS [78]. In an animal model of chronic stress, a decrease of HR was found after stressing mice on a shaking platform for 7 days. This study demonstrated an initial increase of HR in response to the stressor, followed by a transition to lower heart rates during the experiment [10]. As part of an adaptation mechanism under longer-lasting stressful or nociceptive stimulations, increased activity of the parasympathetic branch of the ANS could act as a protective factor, maintaining cardiac adaptability for as long as possible [62].

In humans, studies on HR in chronic pain caused by CIJD provided mixed results. For example, Rensburg and colleagues determined a significantly higher basal HR in women with RA [36], whereas Louthrenoo et al. used a non-invasive cardiovascular reflex test (deep breathing) and found no significant difference in maximum and minimum HR in patients with RA compared with healthy controls [43]. The difference between the maximum and minimum HR, however, was significantly smaller in patients with RA compared to controls. Both studies included RA patients with a relatively short mean disease duration of 4.26 ± 1.2 years [36] and 5.1 ± 3.6 years [43]. By contrast, disease duration in our study was between 1 and 44 years (median: 10 years). It is important to note that there was no significant association between HR under stimulation and pain duration (Figure 3B), suggesting that autonomic control remains intact even in later stages of CIJD.

Similarly, studies on heart rate variability in patients with chronic pain are inconclusive and the selected HRV parameters differ [5]. For example, in 2006, Anichkov and colleagues detected significantly lower HRV in 23 female RA patients compared to matched healthy controls [7]. Contrarily, Vleck and colleagues did not find changes in HRV at rest and under light physical stress in a comparable group (22 female RA patients vs. 15 matched healthy controls) [79].

In our study, we not only measured HR and HRV at rest but also during cognitive stimulation. As expected, HR increased in healthy controls during serial subtraction [6]. Interestingly, our CIJD patients demonstrated a similar increase in HR. So far the most frequently described response to acute stimulation is an HR and LF band increase and a HRV and HF band decrease. This is thought to reflect either a reduction in the parasympathetic or a rise in the sympathetic outflow [38; 78]. In our study, both groups reacted with an increase in HR and SDNN. The CIJD group therefore still seemed to have good cardiac adaptability and reacted appropriately to the stressor. However, only the CIJD reacted with a significant increase in the LF band to cognitive stimulation. The increased LF value in CIJD patients might represent a modified baroreflex [31]. Autonomic imbalance is associated with increased cardiovascular morbidity and mortality [77]. However, to assess the baroreflex more thoroughly was beyond the scope of this study.

### Volumes of the left and right insular cortices

Processing of nociceptive input is a highly complex function of the brain, involving multiple brain regions as well as excitatory and inhibitory mechanisms. Structural MRI studies in chronic pain patients have been inconclusive so far with some showing increased and others, decreased cerebral gray matter density [64; 67]. For this study, we performed an automated analysis of the volumes of the left and right insular cortex using FreeSurfer. In a multiple linear regression model including only CIJD patients with HR at rest as a dependent variable, we found a significant positive relationship with the volume of the left insular cortex. Moreover, we found a significant negative relationship between HR at rest and the volume of the right insula (Table 3).

There is still much debate about the lateralization of insular functions, including autonomic control [50; 55]. A widely acknowledged model of insular lateralization proposes that the right insula predominantly activates sympathetic outflow while the left insula mainly initiates parasympathetic outflow [19; 48]. This model is supported by studies performing unilateral inactivation of the cerebral hemispheres by intracarotid amobarbital procedure [35] and using electrical stimulation of the insulae during epilepsy surgery [51]. Of major clinical importance is the fact that right insular lesions are particularly associated with the development of cardiac arrhythmias [2; 16; 18]. However, this model does not explain all available data and may thus be oversimplified. A recent intraoperative stimulation study did not find differences in HR response between the left and right insula, but between the anterior and posterior parts of both insulae. Left or right posterior insular cortical stimulation predominantly induced tachycardia [15].

Given these controversial findings, the pathomechanistic implications of our results are difficult to assess. Further studies on larger patient populations are needed to replicate these results and to determine if changes in insular volumes might be associated with the increased cardiovascular morbidity and mortality observed in CIJD patients.

ANS changes may not only be caused by central but also by peripheral mechanisms. There is evidence that CIJD can affect afferent peripheral sensory nerves, which in turn lead to sensitization to sympathetic influences [57]. This notion is supported here by a significant increase in sympathetic outflow during stimulation, which could indicate an increased efferent response to the sympathetic nervous system.

### Limitations and future research

One of the major strengths of this study is the matching of patients and controls on an individual basis. Further, this study examined diseases that are based on similar pathophysiology. Nevertheless, further research and longitudinal studies are needed to investigate the influence of chronic pain in patients with rheumatic diseases. There is some evidence that changes in brain areas caused by chronic pain were reversible after pain relief [32]. It is possible that some of the morphological brain changes were diminished due to the plasticity of the brain under consistent pain therapy and pain relief or even occurred in other regions that were not examined in this study. It thus would be interesting to follow newly diagnosed patients with rheumatic diseases before and after drug treatment in a long-term study to compare heart and brain modifications.

Using only cognitive instead of physical stimulation can be critical. This study chose mental stimulation due to its relevance to everyday life. Further, a focus on physical stimulations would have required movements, e.g. handgrip exercise, which in turn would be a risk for creating MRI and pulse data artifacts [42; 52]. In a future study, it would be interesting to examine whether the CIJD group would reach a limit of their heart adaptability earlier than the control group under physical stimulation. Finally, our group size corresponds to other studies that dealt with similar questions or methods [8; 30; 41; 80].

## Conclusion

In summary, our results suggest that HR changes are related to pain duration in CIJD patients. Further, our data established the possibility of insular cortex involvement in the process of ANS changes.

## Data Availability

The datasets generated during the current study are available from the corresponding author on reasonable request.

## Details of authors’ contributions

Study design, ethical approval, and application for funds: CB, PS, MS.

Patient recruitment and data collection: MS, LB.

Data analysis: LB, CB, PS.

Writing manuscript: LB.

Assisting in the review, editing, and writing of the manuscript: CB, PS, MS.

## Acknowledgements

The authors wish to acknowledge the rheumatological clinic of Dr. Markus Voglau, Oldenburg, for the great cooperation in patient recruitment. We thank Katharina Grote and Gülsen Yanç for assisting with MRI data acquisition. We also thank Vivian Baker for editing the final manuscript. The study was supported by the Neuroimaging Unit, University of Oldenburg, funded by grants from the German Research Foundation (DFG; 3T MRI INST 184/152-1 FUGG and MEG INST 184/148-1 FUGG).

